# The Accuracy of Caregiver’s ‘hot to touch’ assessment in paediatric healthcare among children aged 6-35 months with medically-attended diarrhea: Findings from the EFGH-*Shigella* surveillance in Kenya, Malawi, Bangladesh and Peru, 2022-2024

**DOI:** 10.64898/2025.12.16.25342347

**Authors:** Raphael O Anyango, Billy Ogwel, Olivia Schultes, Caren Oreso, Brian O Onyando, Donnie Mategula, Desiree Witte, Wagner Valentino Shapiama Lopez, Pablo Penataro Yori, Taufiqur Rahman Bhuiyan, Syed Qudrat-E-Khuda, Sania Siddiqui, Farah Naz Qamar, Patricia B Pavlinac, Kirkby D. Tickell, Richard Omore

## Abstract

**Introduction:** Confidence in caregivers’ assessment of fever in their children, compared to thermometer readings, could help guide prompt care seeking and appropriate treatment in settings where access to reliable diagnostic tools is limited. Here, we evaluated the accuracy and drivers of caregiver-reported ‘hot-to-touch’ fever compared to digital thermometry among children in the Enterics for Global Health (EFGH) *Shigella* surveillance study.

**Methods:** Children aged 6–35 months with medically attended diarrhea (MAD) enrolled in Kenya, Malawi, Bangladesh, and Peru between August 2022 and August 2024 were included. We trained caregivers to assess and record daily ‘hot-to-touch’ (subjective fever measurement) and digital (thermometer) axillary temperature (fever defined as ≥37.5°C) readings over for 14 days post-enrolment. We calculated site specific and overall accuracy of ‘hot-to-touch’ compared to thermometer-measured fever and used multivariable Poisson regression to identify factors associated with accurate detection.

**Results:** The accuracy of caregiver-reported ‘hot-to-touch’ assessment ranged from 62.1% to72.0% overall and was highest in Bangladesh (83.2%–96.1%) and lowest in Malawi (47.4%–53.4%) over the 14 day assessment period. Accuracy was higher in children with chest indrawing (aPR=1.29, 95% CI: 1.04–1.60) and low respiratory rate (aPR=1.20, 95% CI: 1.11–1.29) and in children from wealthier households (Quintile 5: aPR=1.21, 95% CI: 1.01-1.44). Accuracy was lower among caregivers from households with ≥3 children (aPR=0.88, 95% CI: 0.79–0.99) and for children with low heart rate (aPR=0.76, 95% CI: 0.61–0.94).

**Conclusion:** Suboptimal accuracy of hot-to-touch compared to digital thermometers in detecting fever in this study supports the need for digital thermometer use and context-specific strategies to enhance early detection of fever, particularly in communities living in resource-poor settings.

## Background

Globally, nearly nine percent of an estimated 5.3 million annual preventable child deaths are caused by diarrhea; over 80% of these deaths occur in low- and middle-income countries (LMICs) (1). Fever, an axillary body temperature of ≥ 37.5° Celsius is a common symptom of infection, especially in dehydrating diarrheal illness, and is a major cause of morbidity and mortality in infants and young children world-wide (2). ‘Hot-to-touch’ is a common subjective assessment used by caregivers, especially in LMICs, to assess the presence of fever in their children using a hand to feel for warmth on the child’s forehead, neck or other accessible body parts to sense the degree of heat (3). Although thermometer-measured temperature has been shown to be the gold standard diagnostic tool for detecting fever in young children, in resource poor settings caregiver use of ‘hot-to-touch’ assessment for fever at home is more common (4,5). Household access to thermometers may be limited, with less than 6% of households in Tanzania reported to own a thermometer (6). Recognizing that a caregiver’s early detection of fever at home can increase the likelihood of prompt care seeking, the ‘hot-to-touch’ assessment of fever could be an important child survival strategy in resource-constrained and high disease burden settings.

Few studies have quantitatively evaluated the accuracy of caregiver ‘hot-to-touch’ compared to thermometer-measured fever in infants and young children residing from resource poor settings. The Enterics for Global Health (EFGH) *Shigella* surveillance study was a two-year facility-based study of children aged 6-35 months with medically-attended diarrhea (MAD) to determine *Shigella* incidence in seven countries across Africa, Asia and Latin America (7). We conducted a secondary analysis of data from four study sites to describe the overall accuracy and evaluate the drivers of accurate caregiver ‘hot-to-touch’ assessment of measured fever.

## Methods

### Study Setting and Design

The EFGH-*Shigella* study methods, including the study design and data collection and management procedures, have been described in detail elsewhere (8). We used a prospective population and hospital-based approach to collect fever data from children participating in the EFGH-*Shigella* surveillance from Kenya, Malawi, Bangladesh and Peru study sites, which have been described in detail elsewhere (9–12). In brief, the thermometer sub study involved capturing daily caregiver ‘hot to touch’ assessment and temperature readings of their child over a 14-day period following enrolment with MAD. The thermometer sub-study started recruitment in August, 2022 in the Malawi site.

### Inclusion and Exclusion criteria

All children aged 6-35 months participating in the four EFGH-*Shigella* study sites who had a diarrhea diary completed, with records of both digital axillary thermometer reading and ‘hot-to-touch’ temperature assessments for at least one day post-enrolment, were included.

### Data Collection

All study participants had their household information, demographic, clinical characteristics collected at enrollment. These information were collected through clinician assessment and caregiver reports. As part of the recruitment process, caregivers received training during enrollment on how complete the diarrhea diary including standardizing how to conduct ‘hot to touch’ assessments before taking thermometer readings to reduce bias. Caregivers were counseled to consider the child “hot to touch” by touching the child’s skin, usually on the forehead, neck, or chest, with the back of the hand or fingers. Caregivers were provided with digital thermometers to measure and document temperature daily over the fourteen days following enrolment, to support comparison with ‘hot to touch’ measurements and provided presence or absence of fever [diarrhea diary].

A diarrhea diary is a structured tool that can be adapted to record daily episodes of diarrhea and associated symptoms such as fever in children. Caregivers documented their child’s clinical symptoms including diarrhea duration, stool frequency, hot-to-touch assessment, digital axillary thermometer temperature reading, Oral rehydration solution (ORS), zinc and antibiotic use. Study staff provided training on how to complete the diarrhea diary. Additional details on the completion and collection of diarrhea diary have been provided elsewhere (8).

### Data management and analysis

We defined fever as ≥ 37.5 °C by digital thermometer and ‘hot-to-touch’ was determined by caregiver report in the diarrhea diary. Wealth quintiles were constructed from a weighted sum of country-specific wealth indicators derived from national Demographic Health Surveys (13). The overall and site-specific characteristics of the study population were summarized using frequencies and proportions for categorical variables and median and interquartile range for continuous variables. We calculated the overall and site-specific agreement, sensitivity, specificity, positive predictive value (PPV) and negative predictive value (NPV) of ‘hot to touch’ assessment on thermometer-measured fever by day post-enrollment.

When assessing correlates of ‘hot to touch’ accuracy, we defined the outcome variable as the proportion of daily ‘hot-to-touch’ assessments that matched the thermometer-measured fever designation over a 14-day period, classifying assessments with ≥50% accuracy as having good accuracy. Poisson regression with robust standard errors was utilized to determine factors influencing the accuracy of caregivers’ ‘hot to touch’ assessments as described elsewhere (14). Initially, each potential predictor was examined individually using univariate Poisson regression models. Variables with a p-value ≤0.2 were included in the multivariable model. Both crude and adjusted prevalence ratios (PR) were estimated. We fit models for the combined dataset that included all sites and additionally conducted site-specific analyses to account for potential heterogeneity across the individual country sites from which the results of the adjusted models were reported. We did not run a site-specific model to Peru due to low number of children who had both ‘hot-to-touch’ and temperature readings for at least one day. All analyses were conducted using R version 4.4.1 (R Foundation for Statistical Computing, Vienna, Austria).

### Ethical Considerations

The EFGH protocol was reviewed and approved by the Institutional Review Boards of the individual implementing institutions (9–11,15). Written informed consent was obtained from each caregiver before any research activities were performed.

## Results

### Characteristics of study population

Between August 1^st^, 2022 and August 8^th^, 2024, we screened 9074 children aged 6–35 months presenting with MAD, of whom 2998 (31.2%) were enrolled. Among these, 555 (18.5%) did not have both thermometer and ‘hot-to-touch’ assessments in all the 14-days were excluded, leaving 2443 (81.5%) children for analysis (Figure 1). The median age was 15 months and just over half were male (1333/2442, 54.6%) (Table 1). Socioeconomic status varied by country; most households in Bangladesh and Peru were in higher wealth quintiles, while those in Kenya and Malawi were predominantly from lower quintiles. The median caregiver age was 25 years, with most of them not having any form of employment (1571 [64.4%]). Mild diarrhea by the Modified Vesikari Score (MVS) was most common (1522; 62.3%), with a median duration of 4 days [IQR: 3–6]. The majority of children had normal respiratory rate (2165 [88.6%]) and heart rate (2342 [96.9%]) at enrollment. Additionally, more than a quarter of the children were stunted (632 [26.0%]), while only a few had severe wasting (28 [1.2%]). Nearly a fifth of the children had prolonged diarrhea (415 [17.0%]) with more than two-fifths having a subsequent diarrhea episode (1058 [43.3%]). Variations in clinical characteristics were noted across countries; for example, prolonged diarrhea was highest in Bangladesh (36.5%) and lowest in Kenya (11.1%), while severe diarrhea by MVS (≥ 11) was most frequent in Peru (48%). Respiratory symptoms were uncommon, with 943 (38.6%) reporting cough and 91 (3.7%) diagnosed with pneumonia, predominantly in Malawi (6%). Severe wasting was most prevalent in Bangladesh (2.5%), and stunting was most common in Malawi (28.6%).

**Figure 1.**
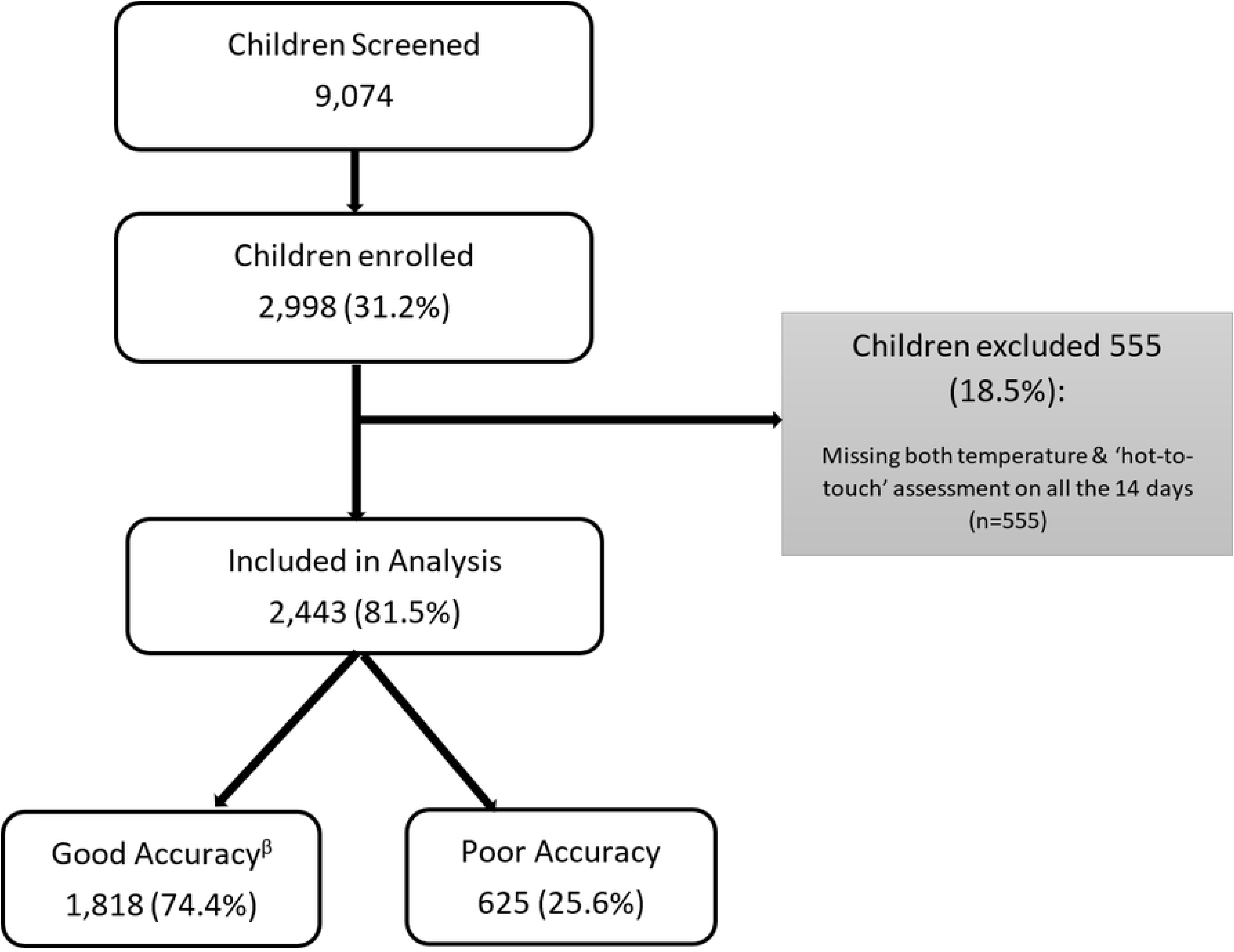
Flowchart of enrolment of children aged 6-35 months presenting with medically attended diarrhea in the thermometer sub-study between September 2023- June 2024. ^β-^Good accuracy- ≥ 50% correct ‘hot-to-touch’ assessments within 14-days post- enrolment; Poor accuracy- < 50% correct ‘hot-to-touch’ assessments within 14-days post- enrolment.

**Table 1.**
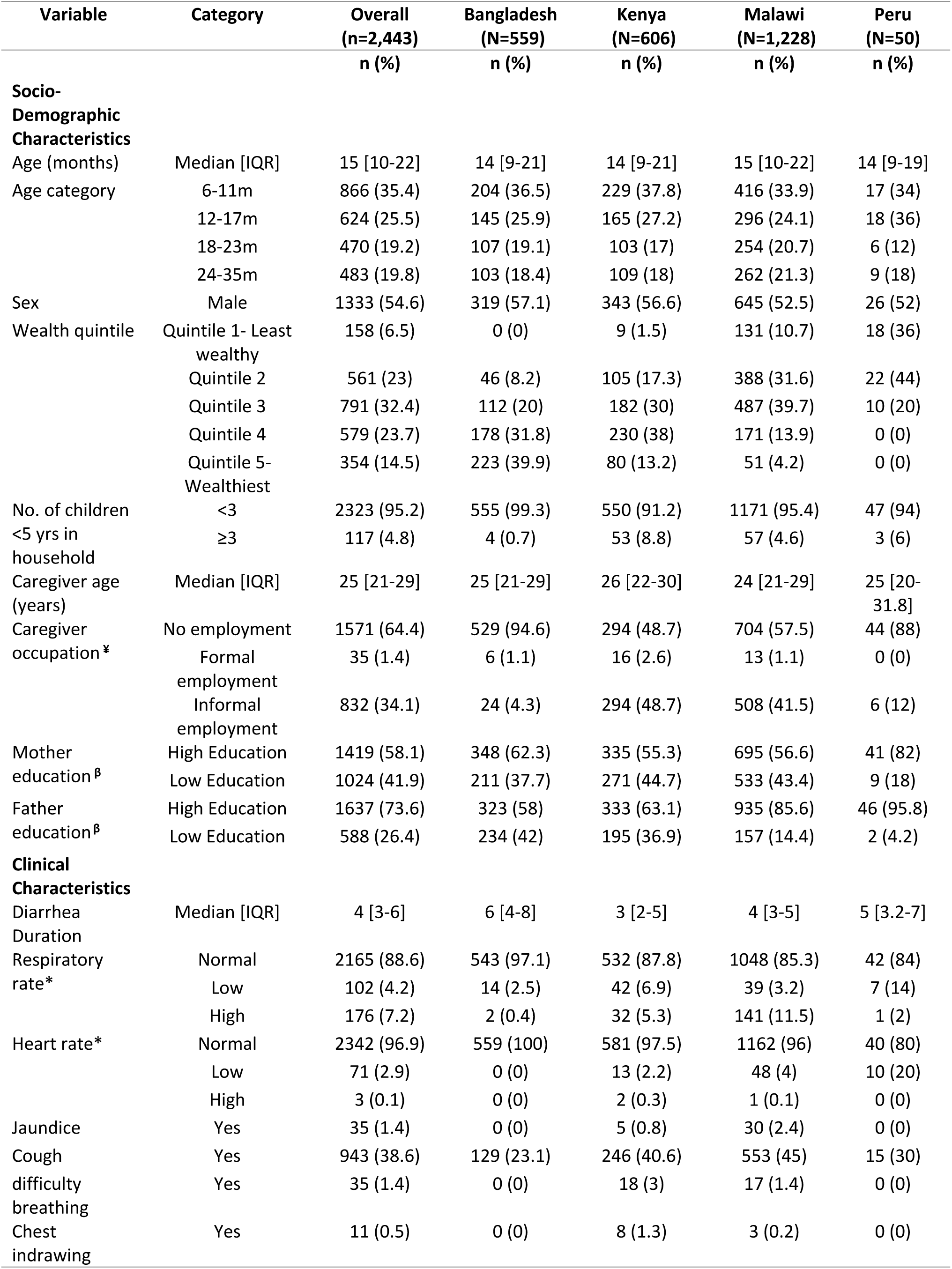
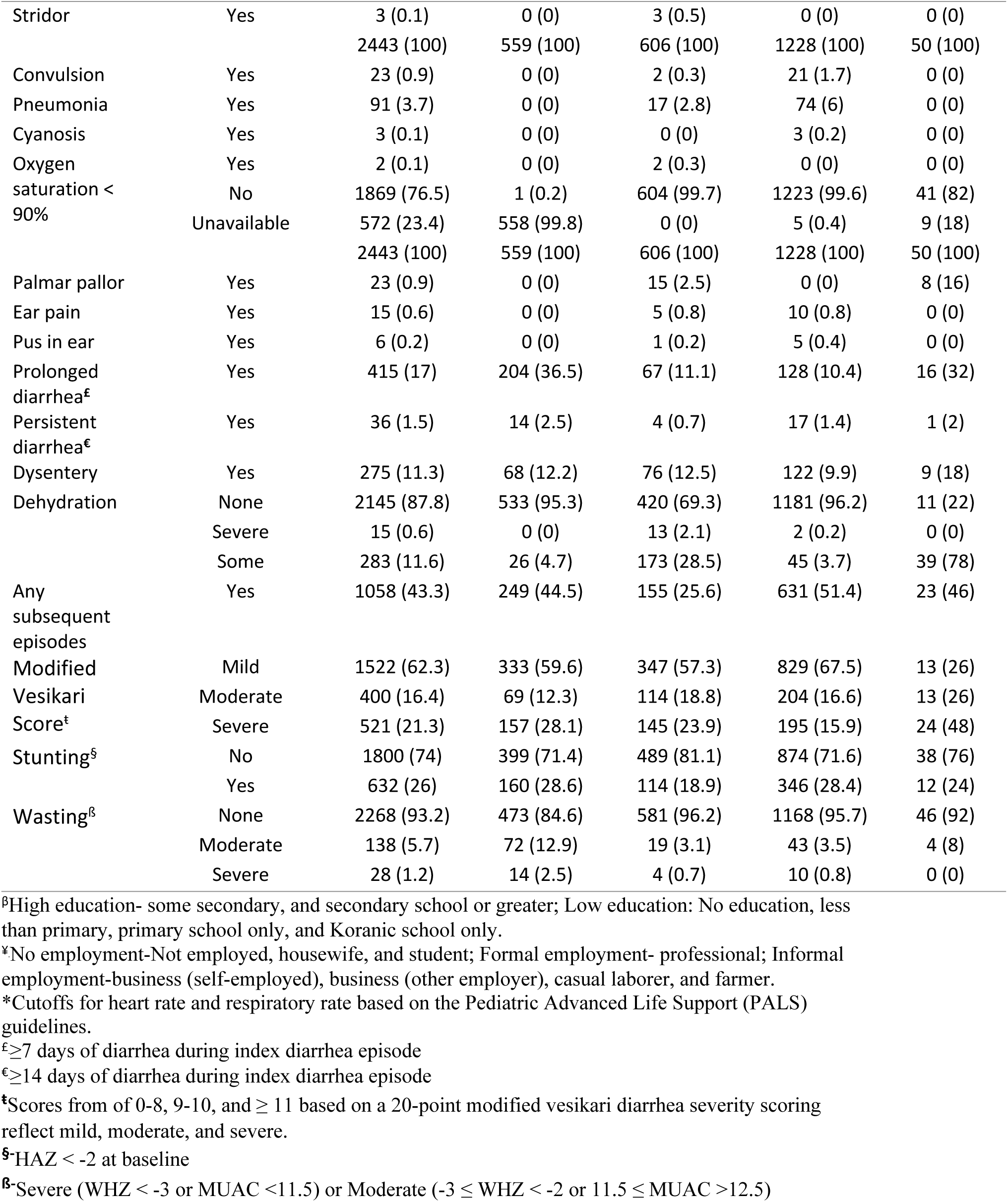
Characteristics of children aged 6-35 months presenting with medically-attended diarrhea enrolled in the thermometer sub-study, 2022-2024

### Performance metrics of caregivers’ ‘Hot-to-Touch’ fever assessment

Based on thermometer readings captured by clinicians, 433 (14.4%) children had fever at the time of enrollment and 1,225 (45.7%) were hot to touch. Among the 81.5% of participants with both temperature and hot-to-touch assessments recorded on at least one day, data completeness ranged from 99.8% on Day 1 to 98.7% on Day 14. This demonstrates the feasibility and acceptability of caregiver-led temperature monitoring using thermometers in this setting. During the fourteen days post-enrollment, thermometer-measured fever prevalence ranged from 7.5- 14.1% and hot to touch ranged from 25.5-42.5%.

The overall accuracy of caregiver accuracy was 74.4%. The accuracy of caregiver reported fever, compared to thermometer, ranged between 62.1%-72.0% by day post-enrollment (Figure 2). Sensitivity decreased from 68.1% on day 1 and to 43.4% on day 14, while specificity increased from 61.1% on day 1 to 74.0% on day 14. There were site differences in the accuracy of caregivers ‘hot-to-touch’ assessment by day post-enrollment, with caregivers in Bangladesh having the highest accuracy (83.2%-96.1%) and those in Malawi having the lowest accuracy (47.4%-53.4%) (Figure 3). The performance metrics stratified by site are shown in Figure 4.

**Figure 2:**
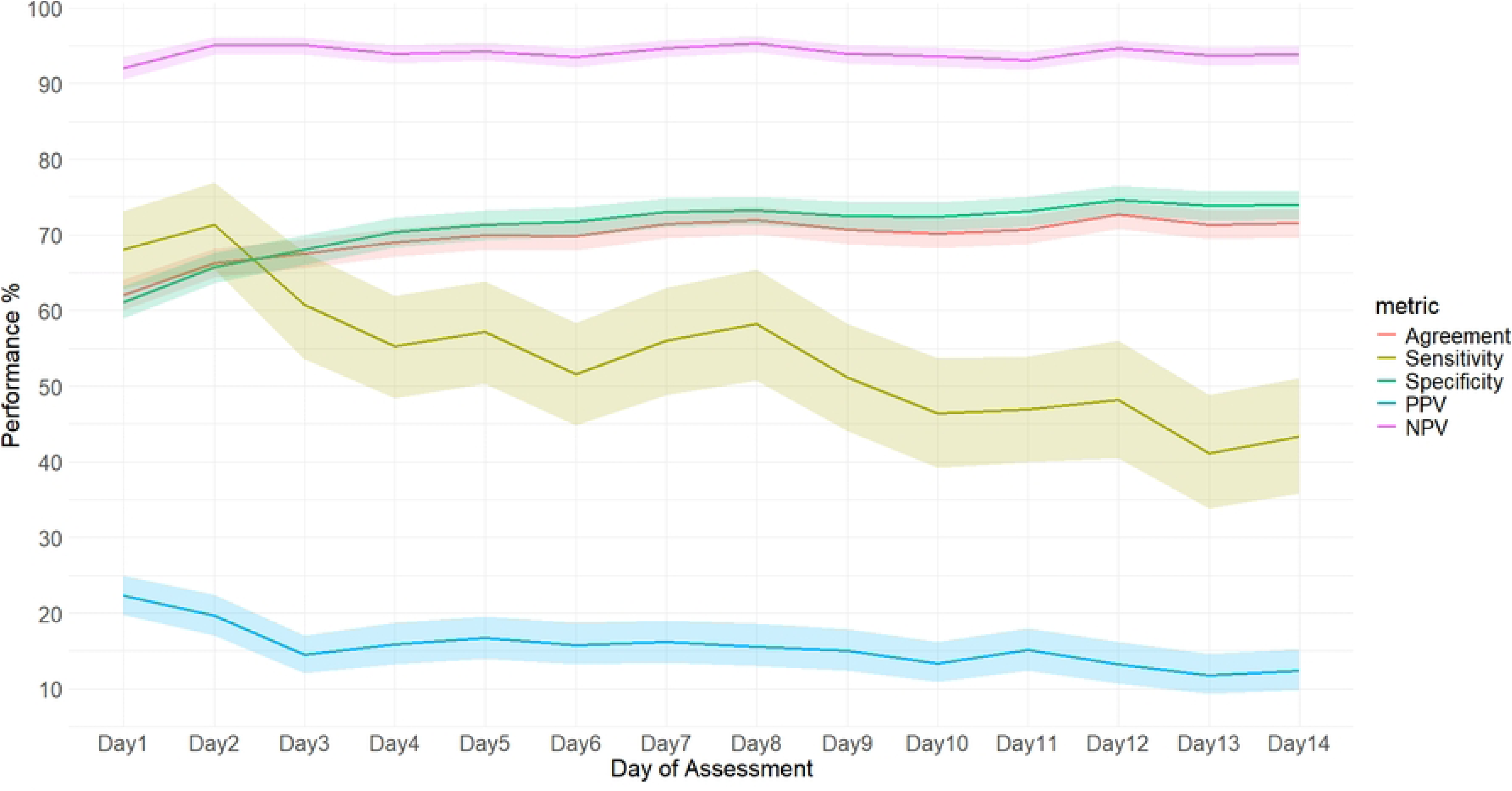
Performance metrics of caregivers’ ‘hot-to-touch’ assessment of fever in all study sites combined: 2022-2024. *PPV- Positive predictive value; NPV- Negative predictive value

**Figure 3:**
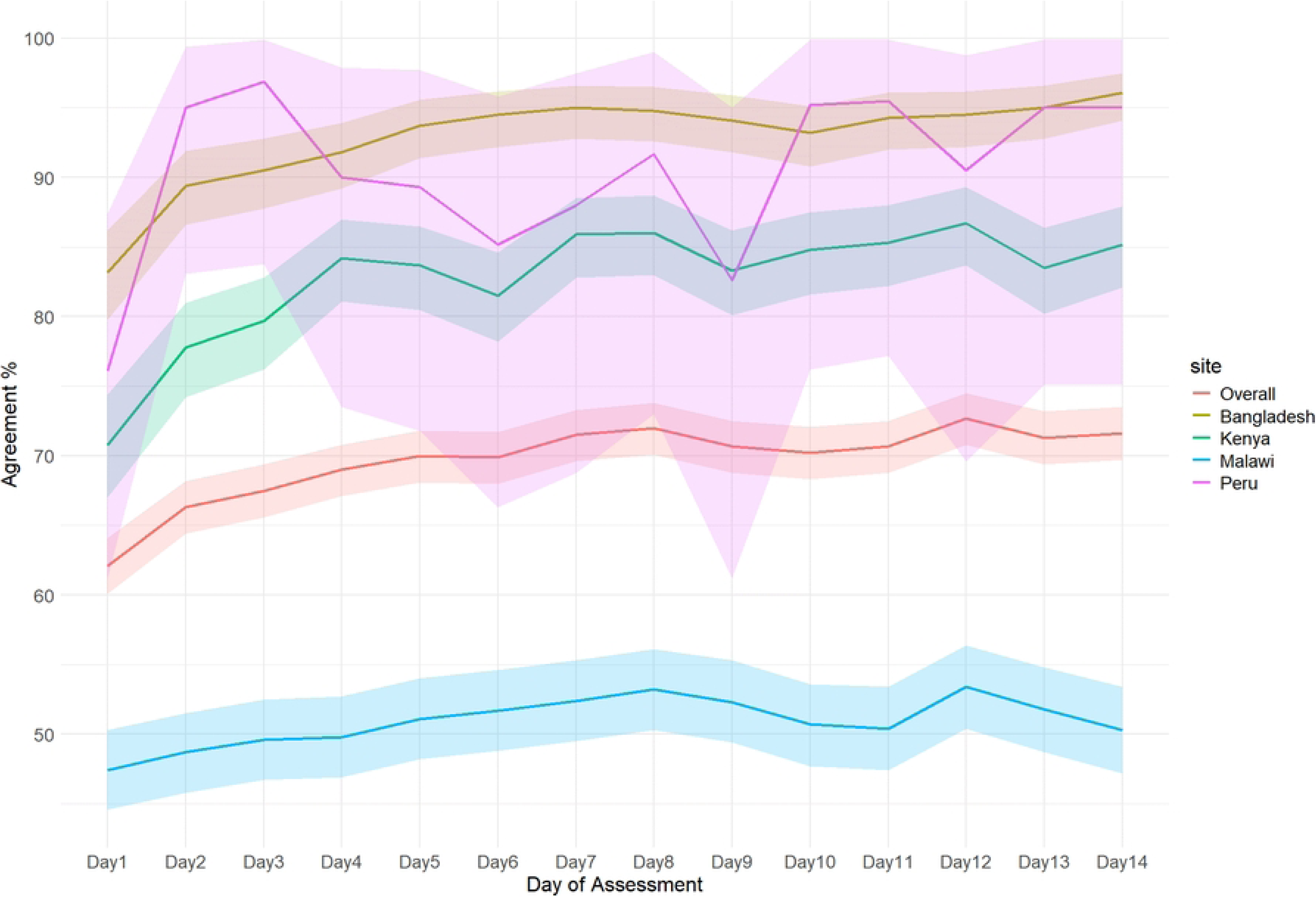
Accuracy of caregivers’ ‘hot-to-touch’ assessment of fever across study sites: 2022-2024.

**Figure 4.**
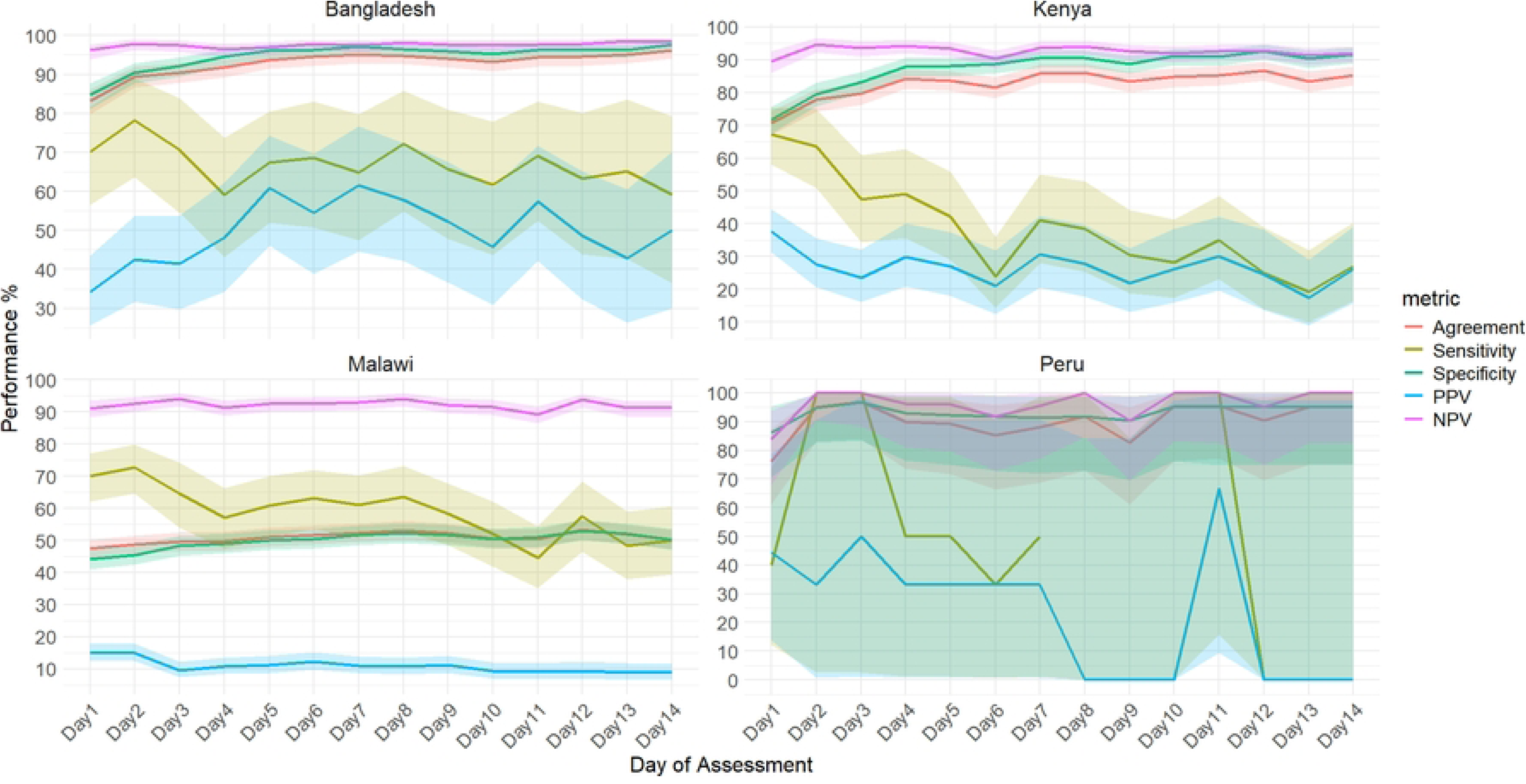
Assessment of performance metrics of caregivers’ ‘hot-to-touch’ assessment across study sites: 2022-2024. *PPV- Positive predictive value; NPV- Negative predictive value

### Factors Associated with the caregiver’s accuracy of ‘hot to touch’ assessment of fever

In overall univariate models, sex, wealth quintile, father education, chest indrawing, palmar pallor, prolonged diarrhea, diarrheal severity and wasting were associated with higher caregiver accuracy in the ‘hot to touch’ assessment of fever. Conversely, jaundice, cough, and subsequent diarrheal episodes were associated with lower caregiver accuracy.

In the multivariate model, caregivers of children who presented with chest indrawing and low respiratory rate had 29%, aPR=1.29, 95% CI: 1.04–1.60) and 20% (aPR=1.20, 95% CI: 1.11–1.29) higher accuracy in the subjective assessment of fever, respectively, compared to those who did not (Table 2). Additionally, compared to caregivers from households with lowest wealth index, those from households with a higher wealth status had higher accuracy (quintile 3: aPR=1.21, 95% CI: 1.02–1.44; quintile 4: aPR=1.22, 95% CI: 1.02–1.45; quintile 5: aPR=1.21, 95% CI: 1.01–1.44). However, compared to caregivers from Bangladesh, the accuracy of those from Malawi was lower by 43% (aPR=0.57, 95% CI: 0.53–0.61). Similarly, caregiver accuracy was 12% lower in households with three or more children (aPR=0.88, 95% CI: 0.79–0.99) and 24% lower among children with low heart rate (aPR=0.76, 95% CI: 0.61–0.94), compared to caregivers from households with fewer than three children and those whose children had normal heart rate, respectively. Factors like caregiver occupation, prolonged diarrhea, and diarrhea severity did not retain significance in the final multivariate model.

**Table 2.**
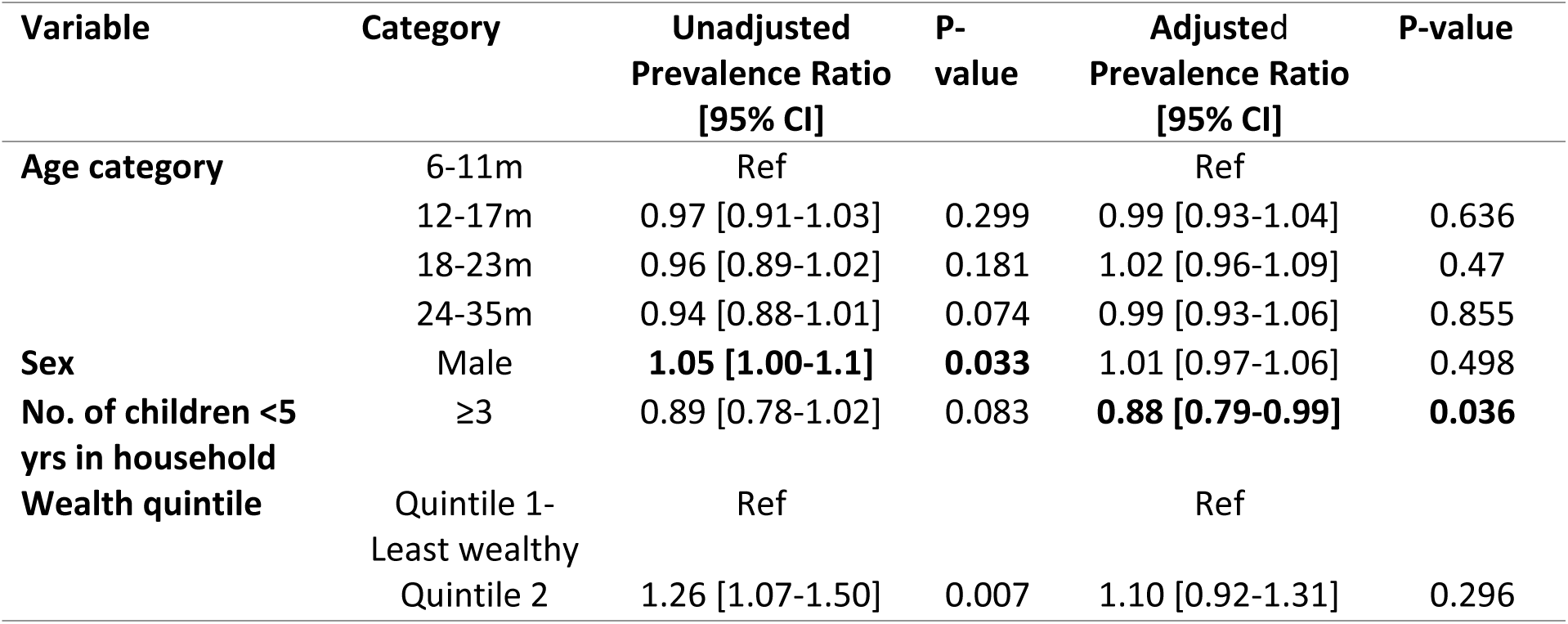
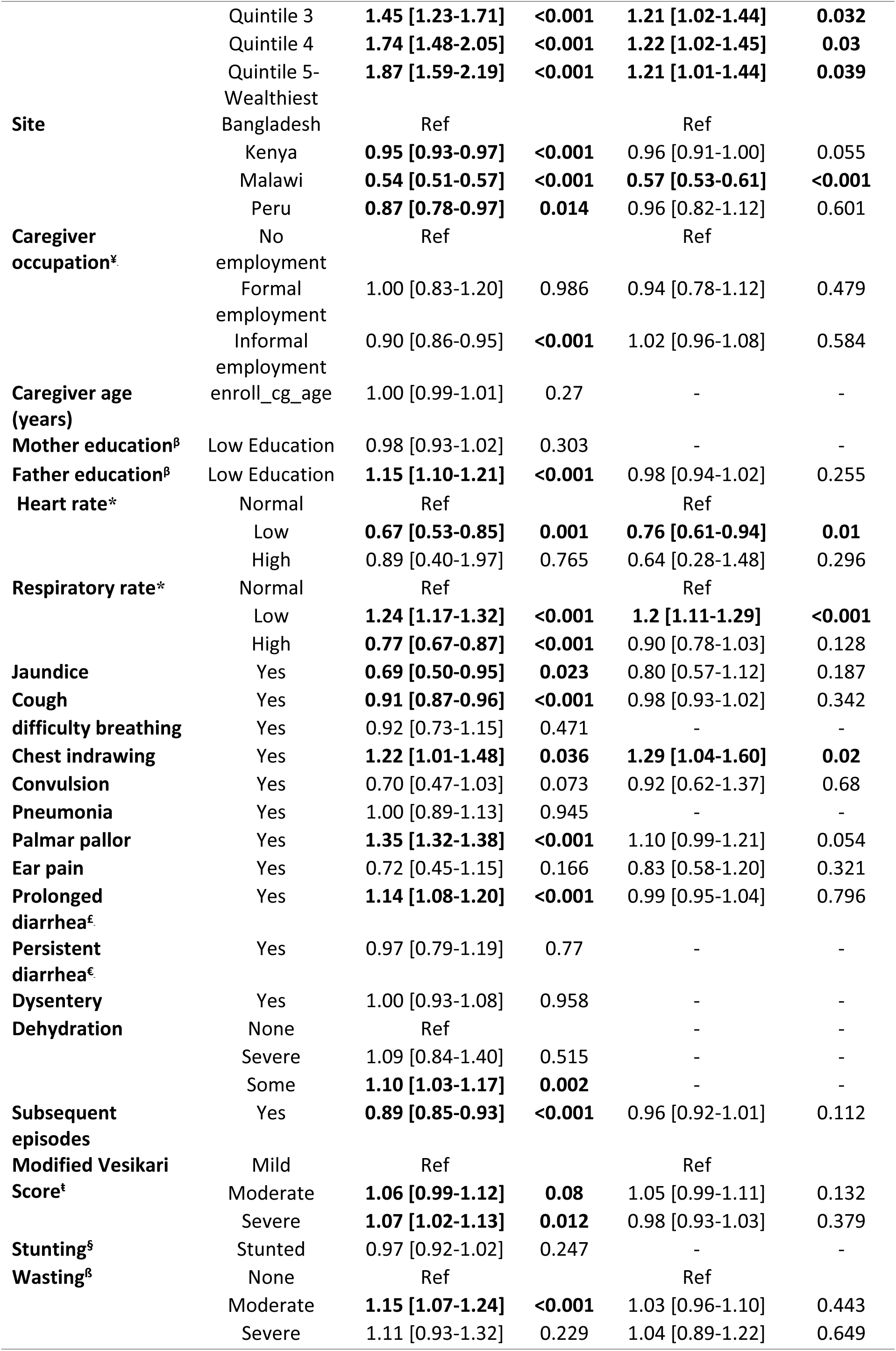
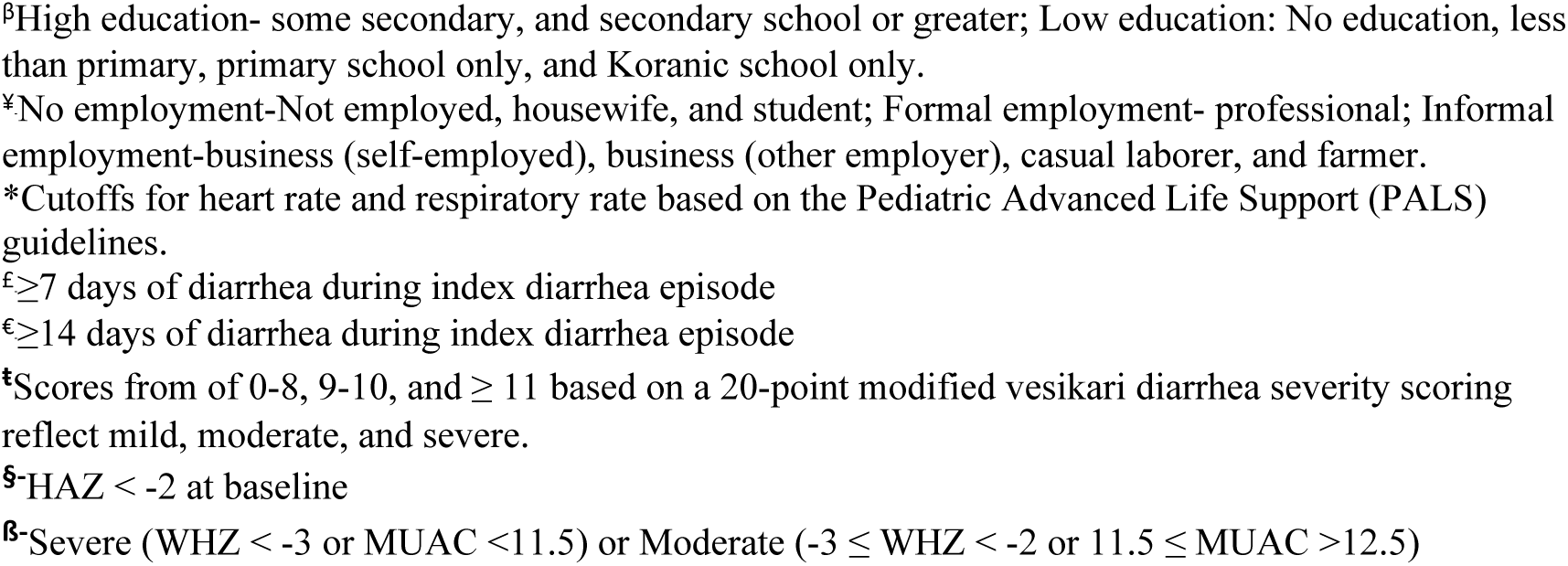
Factors associated with accuracy of caretakers’ tactile assessment of fever among children aged 6-35 months presenting with medically-attended diarrhea, 2022-2024

From the site-specific models, we observed distinct factors to be associated with the accuracy of caregivers’ ‘hot to touch’ assessment of fever among children aged 6–35 months presenting with MAD in each of the sites (Bangladesh, Kenya, and Malawi). In Bangladesh, moderate diarrhea severity (aPR = 1.02, 95% CI: 1.00–1.03) was the only factor significantly associated with assessment accuracy (Table 3). In Kenya, factors such as low heart rate (aPR = 1.09, 95% CI: 1.03–1.14), high respiratory rate (aPR = 1.08, 95% CI: 1.03–1.12), jaundice (aPR = 1.06, 95% CI: 1.01–1.11), difficulty breathing (aPR = 1.15, 95% CI: 1.03–1.28), ear pain (aPR = 1.08, 95% CI: 1.03–1.15), persistent diarrhea (aPR = 1.09, 95% CI: 1.04–1.16), and stunting (aPR = 1.06, 95% CI: 1.01–1.11) were all associated with higher accuracy, while male sex (aPR = 0.95, 95% CI: 0.91–0.99) was associated with lower accuracy. In Malawi, increased caregiver assessment accuracy was associated with low respiratory rate (aPR = 1.69, 95% CI: 1.45–1.96), chest indrawing (aPR = 1.99, 95% CI: 1.33–2.98), and pneumonia (aPR = 1.28, 95% CI: 1.07–1.53), while lower accuracy was observed among children with low heart rate (aPR = 0.50, 95% CI: 0.32–0.79) and in households with three or more children (aPR = 0.65, 95% CI: 0.43–0.97) (Table 3).

**Table 3.** Factors associated with accuracy of caretakers’ tactile assessment of fever among children aged 6-35 months presenting with medically-attended diarrhea stratified by Country-Site, 2022-2024

| Variable | Category | Bangladesh | Kenya | Malawi |
| --- | --- | --- | --- | --- |
|  |  | Adjusted<br>Prevalence Ratio<br>[95% CI] | Adjusted<br>Prevalence Ratio<br>[95% CI] | Adjusted<br>Prevalence Ratio<br>[95% CI] |
| Age category | 6-11m | Ref | Ref | Ref |
|  | 12-17m | 1.00 [0.97-1.02] | 1.02 [0.95-1.08] | 0.95 [0.81-1.11] |
|  | 18-23m | 1.01 [0.99-1.03] | 1.02 [0.95-1.09] | 1.02 [0.87-1.19] |
|  | 24-35m | 0.99 [0.96-1.02] | 1.04 [0.98-1.11] | 0.96 [0.81-1.13] |
| Sex | Male | 1.01 [0.99-1.03] | **0.95 [0.91-0.99]** | 1.07 [0.96-1.20] |
| No. of children <5 yrs in household | ≥3 | 1.02 [0.99-1.04] | - | **0.65 [0.43-0.97]** |
| Wealth quintile | Quintile 1-Least<br>wealthy | Ref |  | Ref |
|  | Quintile 2 | - | - | 1.06 [0.84-1.34] |
|  | Quintile 3 | 0.97 [0.94-1.00] | - | 1.21 [0.97-1.52] |
|  | Quintile 4 | 0.99 [0.97-1.00] | - | 1.25 [0.98-1.60] |
|  | Quintile 5-Wealthiest | 0.99 [0.98-1.01] | - | 1.25 [0.90-1.72] |
| Caregiver occupation<sup>‡</sup> | No<br>employment | Ref |  |  |
|  | Formal<br>employment | 1.02 [0.99-1.04] | - | - |
|  | Informal<br>employment | 1.02 [0.99-1.05] | - | - |
| Mother education<sup>§</sup> | Low<br>Education | - | - | - |
| Father education<sup>§</sup> | Low<br>Education | - | 1.03 [0.99-1.07] | **0.8 [0.67-0.97]** |
| Heart rate* | Normal |  | Ref |  |
|  | Low |  | **1.09 [1.03-1.14]** | **0.5 [0.32-0.79]** |
|  | High |  | 0.96 [0.89-1.04] | - |
| Respiratory rate* | Normal | Ref | Ref | Ref |
|  | Low | 1.02 [0.99-1.04] | 1.05 [0.99-1.12] | **1.69 [1.45-1.96]** |
|  | High | 1.00 [0.99-1.01] | **1.08 [1.03-1.12]** | 0.82 [0.67-1.01] |
| Jaundice | Yes | - | **1.06 [1.01-1.11]** | 0.79 [0.49-1.29] |
| Cough | Yes | - | - | - |
| difficulty breathing | Yes | - | **1.15 [1.03-1.28]** | 0.69 [0.34-1.44] |
| Chest indrawing | Yes | - | 0.92 [0.81-1.04] | **1.99 [1.33-2.98]** |
| Convulsion | Yes | - | 1.06 [0.95-1.18] | - |
| Pneumonia | Yes | - | 1.03 [0.97-1.09] | **1.28 [1.07-1.53]** |
| Palmar pallor | Yes | - | 1.04 [0.99-1.09] | - |
| **Ear pain** | Yes | - | **1.08 [1.03-1.15]** | - |
| **Prolonged diarrhea<sup>£</sup>** | Yes | - | - | - |
| **Persistent diarrhea<sup>£</sup>** | Yes | - | **1.09 [1.04-1.16]** | - |
| **Dysentery** | Yes | - | - | - |
| **Dehydration** | None | - | - | - |
|  | Severe | - | - | - |
|  | Some | - | - | - |
| **Subsequent episodes** | Yes | - | 0.97 [0.92-1.03] | 0.9 [0.81-1.01] |
| **Modified Vesikari Score<sup>‡</sup>** | Mild | - | Ref | Ref |
|  | Moderate | **1.02 [1.00-1.03]** | 0.95 [0.89-1.02] | **1.15 [1.00-1.32]** |
|  | Severe | 1.01 [0.99-1.03] | 0.99 [0.94-1.05] | 0.89 [0.75-1.05] |
| **Stunting<sup>§</sup>** | Stunted | - | 1.06 [1.01-1.11] | - |
| **Wasting<sup>§</sup>** | None | - | Ref | - |
|  | Moderate | 1.00 [0.97-1.03] | 1.02 [0.98-1.07] | - |
|  | Severe | 1.01 [0.99-1.02] | 0.65 [0.30-1.42] | - |

<sup>β</sup>High education- some secondary, and secondary school or greater; Low education: No education, less than primary, primary school only, and Koranic school only.
<sup>¥</sup>No employment-Not employed, housewife, and student; Formal employment- professional; Informal employment-business (self-employed), business (other employer), casual laborer, and farmer.
<sup>\*</sup>Cutoffs for heart rate and respiratory rate based on the Pediatric Advanced Life Support (PALS) guidelines.
<sup>£</sup>≥7 days of diarrhea during index diarrhea episode
<sup>€</sup>≥14 days of diarrhea during index diarrhea episode
<sup>‡</sup>Scores from 0-8, 9-10, and ≥ 11 based on a 20-point modified vesikari diarrhea severity scoring reflect mild, moderate, and severe.
<sup>§</sup>-HAZ < -2 at baseline
<sup>β</sup>-Severe (WHZ < -3 or MUAC <11.5) or Moderate (-3 ≤ WHZ < -2 or 11.5 ≤ MUAC >12.5)

## Discussion

Despite the widespread use of ‘hot-to-touch’ assessments as a first line of fever detection by caregivers in LMICs, its diagnostic accuracy compared to thermometer-measured fever is poorly characterized. We evaluated the accuracy and drivers of caregiver-reported ‘hot-to-touch’ fever compared to thermometer-measured fever among children aged 6–35 months with MAD in four resource-limited settings. We observed that hot to touch may substantially overestimate fever, sometimes by more than three-fold, when compared to thermometer-measured fever, aligning with similar observations reported by Felix et al. (2). We also demonstrated the feasibility and acceptability of caregiver-led temperature monitoring using thermometers, with more than four-fifths of caregivers recording both temperature and hot-to-touch assessments on at least one day. Data completeness in this group remained high throughout the follow-up period (≥ 98.7%).

The accuracy of caregivers’ ‘hot-to-touch’ assessments by day post-enrollment were moderate (62%-72%) and varied considerably across countries, with the highest accuracy observed in Bangladesh (83%-96%) and the lowest in Malawi (47%-53%). Accuracy declined over the days following enrollment, with sensitivity decreasing and specificity increasing during the 14-day follow-up period. Caregiver ‘hot-to-touch’ accuracy was higher among caregivers from wealthier households and for children presenting without signs of severe respiratory distress, including chest indrawing or a high respiratory rate, while accuracy was lower in households with three or more children under five, as well as among children presenting with lower heart rates. This highlights the need for continued use of conventional fever detection methods in clinical settings.

Our finding of the moderate accuracy of caregivers’ ‘hot-to-touch’ assessments aligns with previous studies indicating that such subjective evaluations are often unreliable for detecting fever (5,16,17). This highlights the need to equip community health workers who are closer to households with thermometers and reinforces the continued use of conventional, objective fever detection methods in clinical settings. Furthermore, the observed heterogeneity in caregivers’ ‘hot-to-touch’ assessments across sites may be attributed to cultural differences, healthcare-seeking behaviors, or environmental factors influencing caregivers’ perceptions of fever (18). In regions with endemic malaria and infectious disease, caregivers might be more alert and unintentionally overestimate fever as a symptom of potential infection (19) . In settings like Malawi, where accuracy of caregivers’ ‘hot-to-touch’ assessments was low, there is need for enhanced awareness of healthcare seeking strategies targeted at child survival. In contrast, in Bangladesh where accuracy was much higher, leveraging health care seeking awareness on fever detection by ‘hot-to-touch’ could improve prompt treatment in such settings. In both settings, there was no concern with use of digital thermometers in homes and thus thermometers could be used. These findings highlight the need to consider site-specific factors when designing public health interventions such as fever detection and management strategies.

Lower respiratory rates, chest indrawing and higher wealth quintiles were associated with improved caregivers’ accuracy in assessing fever. The latter association may be linked to better overall health knowledge, access to healthcare resources, and socioeconomic advantages that influence caregiving practices. Higher wealth quintiles suggest economic empowerment and access to better healthcare, which can enhance caregiver ability to assess symptoms and seek care promptly, consistent with findings from previous studies (20). In parallel, it has been documented that fever may by itself alter respiratory rate, this could in part explain why chest indrawing and lower respiratory rates could potentially make febrile symptoms easier to identify (21). This finding emphasizes the role of overall child health in accurate fever assessment. Taken together, these findings highlight the need of addressing socioeconomic disparities and promoting health education to improve caregivers’ accuracy in fever detection to enhance child survival. Public health strategies should focus on empowering caregivers in lower socioeconomic settings through health education and access to affordable healthcare tools, fostering equitable health outcomes and timely illness detection.

Having three or more children under five years in a household and a lower heart rate were associated with reduced caregiver ‘hot-to-touch’ accuracy. A higher number of children in households could reduce accuracy due to a potential overload on caregivers, which may hinder their ability to assess individual children’s fever accurately. This is congruent with a World Health Organization systematic review found that a caregiver’s ability to appropriately respond to a child’s signals depends on their overall sensitivity; that is, the caregiver’s capacity to notice, understand, and respond consistently to the child’s needs—a quality that is diminished when caring for several children under five (22). Moreover, a low heart rate is an indicator of severe illness, which may be overlooked by caregivers using only ‘hot-to-touch’ fever assessment. This observation is corroborated by a systematic review, which found that in resource-limited settings, caregiver recognition of syndromes associated with fever—such as malaria, pneumonia, and diarrhea—is generally poor (18). Public health initiatives should focus on providing caregivers with practical training and support, particularly in high-burden households, to enhance their skills in fever assessment and ensure timely medical intervention for at-risk children.

A key limitation is that clinical factors were recorded only at enrollment, meaning the analysis could not account for any changes in the child’s condition during the 14-day follow-up. Another underlying assumption is that the characteristics of children included in the study are comparable to those who had diarrhea but either did not seek care or sought care at non-sentinel health facilities. However, this may not hold true, highlighting the need for caution when generalizing the findings.

## Conclusion

While ubiquitous in LMICs, the ‘hot-to-touch’ fever assessment exhibits only moderate accuracy compared to digital thermometers, is highly variable across settings, and often overestimates fever prevalence. This underscores the method’s inherent limitations and necessitates a dual public health approach. First, it is critical to maintain thermometers as the standard in clinical settings while expanding their availability to community health workers. Second, public health interventions to bolster prompt detection of fever should be context-specific in addressing the root causes of inaccuracy through targeted health education and socioeconomic support, thereby creating a more robust safety net for at-risk children.

## Data Availability

The deidentified and anonymized EFGH datasets, data dictionaries, statistical analysis plan, CRFs and study protocol were made publicly available on the Vivli repository in December 2025. Access to the data and supporting documents is available on request at Vivli DOI - PR00011860 and requires the execution of a Data Use Agreement

## Acknowledgements

The authors extend their deep gratitude to the families who participated in the study, and to the clinical and field teams whose dedication and hard work made this research possible. We are also grateful to the physicians, administrators, and Ministry of Health officials at each study site for their generous provision of facilities, operational support, and gatekeeper permissions that enabled the successful implementation of the study. We thank the EFGH consortium and the Nyanja Health Institute who facilitated and contributed to the Manuscript Writing cohort program including Beth Tippett Barr and Erika Feutz for their support and guidance. Special thanks to the EFGH Consortium—including the University of Washington, University of Virginia, University of Maryland Baltimore, and Emory University—for their technical guidance and oversight.

## Data Availability

The EFGH statistical analysis plan (https://clinicaltrials.gov/study/NCT06047821) and study protocol (https://academic.oup.com/ofid/issue/11/Supplement_1) were made publicly available. The datasets were deidentified and anonymized and will be publicly available upon publication of the manuscript.

